# Shared genetic etiology between chronic diseases and heart failure risk: the dual role of leukocyte telomere length

**DOI:** 10.1101/2023.08.03.23293621

**Authors:** Jason Y.Y. Wong, Batel Blechter, Zhonghua Liu, Jianxin Shi, Véronique L. Roger

## Abstract

**Introduction:** Genetic susceptibility to various chronic diseases has been shown to influence heart failure (HF) risk. However, the pathogenic mechanisms underlying these associations, particularly the role of phenotypic leukocyte telomere length (LTL), is unknown. We investigated the shared genetic etiology between chronic diseases, various traits, and HF risk, and whether LTL mediates or modifies these relationships.

**Methods:** We conducted prospective cohort analyses on 404,883 European participants from the UK Biobank, including 9,989 incident HF cases. Multivariable Cox regression was used to estimate associations between HF risk and 24 polygenic risk scores (PRSs) for various diseases or traits previously generated in the UK Biobank using a Bayesian approach. We assessed multiplicative interactions between the PRSs and LTL previously measured in the UK Biobank using quantitative PCR. Mediation analyses were conducted to estimate the proportion of the total effect of PRSs acting indirectly through LTL, an integrative marker of biological aging.

**Results:** We identified 9 PRSs associated with HF risk, including those for various cardiovascular diseases or traits, rheumatoid arthritis (P=1.3E-04), and asthma (P=1.8E-08). Additionally, longer LTL was strongly associated with decreased HF risk (P-trend=1.7E-08). Notably, the asthma PRS had a super-multiplicative interaction with LTL (P-interaction=2.8E-03). However, LTL mediated only 1.13% (P<0.001) of the total effect of the asthma PRS on HF risk.

**Conclusions:** Our findings shed light onto the shared genetic etiology between HF risk, asthma, rheumatoid arthritis, and other traits. Longer LTL strengthened the genetic effect of asthma on HF, supporting their utility in risk stratification analyses.

## Introduction

Heart failure (HF) is a phenotypically complex syndrome that is commonly characterized by the diminished ability of the ventricle to pump or fill with blood ^1–4^. HF is a downstream consequence of most cardiovascular diseases (CVD) including coronary heart disease, high blood pressure, atrial fibrillation (AF), and cardiomyopathies ^5^. Genome-wide association studies (GWAS) have found a modest contribution of genetic susceptibility to HF risk, with heritability estimates ranging from 8.8% ^6^ to 26% ^7^. Thus far, 12 independent genome-wide significant variants have been identified to be associated with HF among European populations ^6, 8^. All of the identified genetic variants had associations with coronary artery disease (CAD), AF, or reduced left ventricular function, which strongly supports the downstream nature of HF relative to CVD ^6, 8^. Besides CVD, recent studies have shed light on bi-directional relationships between HF development and various cancers ^9–12^, which may be attributed in part to shared biologic pathways ^13, 14^. A prominent molecular pathway for the initiation and progression of both HF and cancers is increased inflammatory states or dysfunction ^15^, including those due to autoimmune diseases such as rheumatoid arthritis ^16, 17^. Taken together, these observations suggest shared genetic susceptibility across CVDs, cancers, various inflammatory phenotypes and the pathogenesis of heart failure. However, the biological processes underlying these relationships are unclear. Given that chronological age is a strong risk factor for heart failure (as well as other chronic diseases), genetic susceptibility potentially influences heart failure risk through accelerated biological aging or weathering. One such biomarker reflective of biological aging is leukocyte telomere length (LTL).

Telomeres are hexameric (TTAGGG)_n_ repeats at the ends of linear chromosomes that protect the genome from progressive degradation after each round of mitosis. LTL has a modest genetic heritability of approximately 8.1% ^18, 19^, with the remaining variation explained largely by potential missing heritability, non-genetic environmental factors, and technical variables. LTL has an apparent dual-nature and reflects two broad components. First, LTL reflects exogenous environmental exposures that cause genotoxic damage to the DNA of hematopoietic stem cells, their derivative leukocytes, as well as highly replicative target tissues ^20^. Second, LTL reflects endogenous biological pathways related to biological aging or weathering ^21–30^. Taken together, LTL can be considered as an integrative biomarker of genomic instability that captures the cumulative burden of biological stressors across the lifespan. Shorter phenotypic LTL, which reflects a higher load of biological stressors, has been consistently found to be associated with increased risk of HF ^18, 31^, CVD ^32^, and coronary heart disease^33^. Additionally, measured or genetically-determined LTL have been reported to have relationships of various directions with different cancers ^34–40^ as well as inflammatory conditions ^41^. However, the precise role of LTL in the pathogenic mechanism underlying the relationship between genetic susceptibility and HF risk is unknown.

To investigate these gaps in knowledge, we leveraged the extensive questionnaire, clinical, and molecular/genetic data from the UK Biobank, a prospective cohort study of nearly half a million adults residing in the United Kingdom. The UK Biobank is among the premier cohort studies for the investigation of genetic etiology among European populations and is the single largest resource for human telomere length data in the world ^18, 19^. Our overarching goal was to gain mechanistic insight into the role of phenotypic LTL in influencing the effect of genetic susceptibility on HF risk. Specifically, we were interested in: 1) agnostically exploring the effect of genetic susceptibility to cardiovascular diseases or traits, cancers, metabolic diseases, and inflammatory conditions on HF risk; 2) investigating whether LTL, particularly the environmentally-determined components, modifies the strength of associations between genetic susceptibility and HF risk; and 3) estimating the degree in which LTL, particularly the biological aging component, mediates the total effect of genetic susceptibility on HF risk. Our findings improve the understanding of the shared biological mechanisms underlying HF and other non-malignant and malignant diseases, as well as support the use of LTL and genetics in risk stratification analyses to identify high-risk subgroups that could benefit from early intervention.

## Materials and Methods

The UK Biobank dataset used for this study is publicly available at: https://bbams.ndph.ox.ac.uk/ams/. Our analyses were conducted under UK Biobank project number: 28072 (datasets from August 2022 – June 2023). Additional metadata and codes are available from the corresponding author upon reasonable request. The detailed methods are available within the *Online Supplement*. Briefly, we used a multi-step approach to investigate the interrelationships between genetic susceptibility to various diseases or traits, LTL, and HF risk (Figure 1). Among participants of European ancestry, we first agnostically assessed the prospective associations between 24 previously derived CVD-, cancer-, and inflammation-related polygenic risk scores (PRSs) ^42^ and future risk of developing HF. Second, among the PRSs identified to be significantly associated with HF risk, we then investigated whether the PRSs statistically interact with measured LTL to influence HF risk. Among PRSs with evidence of multiplicative interaction with LTL, we conducted further analyses stratified by quartiles of LTL. In parallel, we agnostically assessed associations between the 24 PRSs and LTL measured at the time of study enrollment. Here, we qualitatively looked for overlapping findings with the PRS-HF analyses. Lastly, we determined whether the effect of the significant PRSs on HF risk was mediated through biological aging processes reflected by altered LTL.

**Figure 1:**
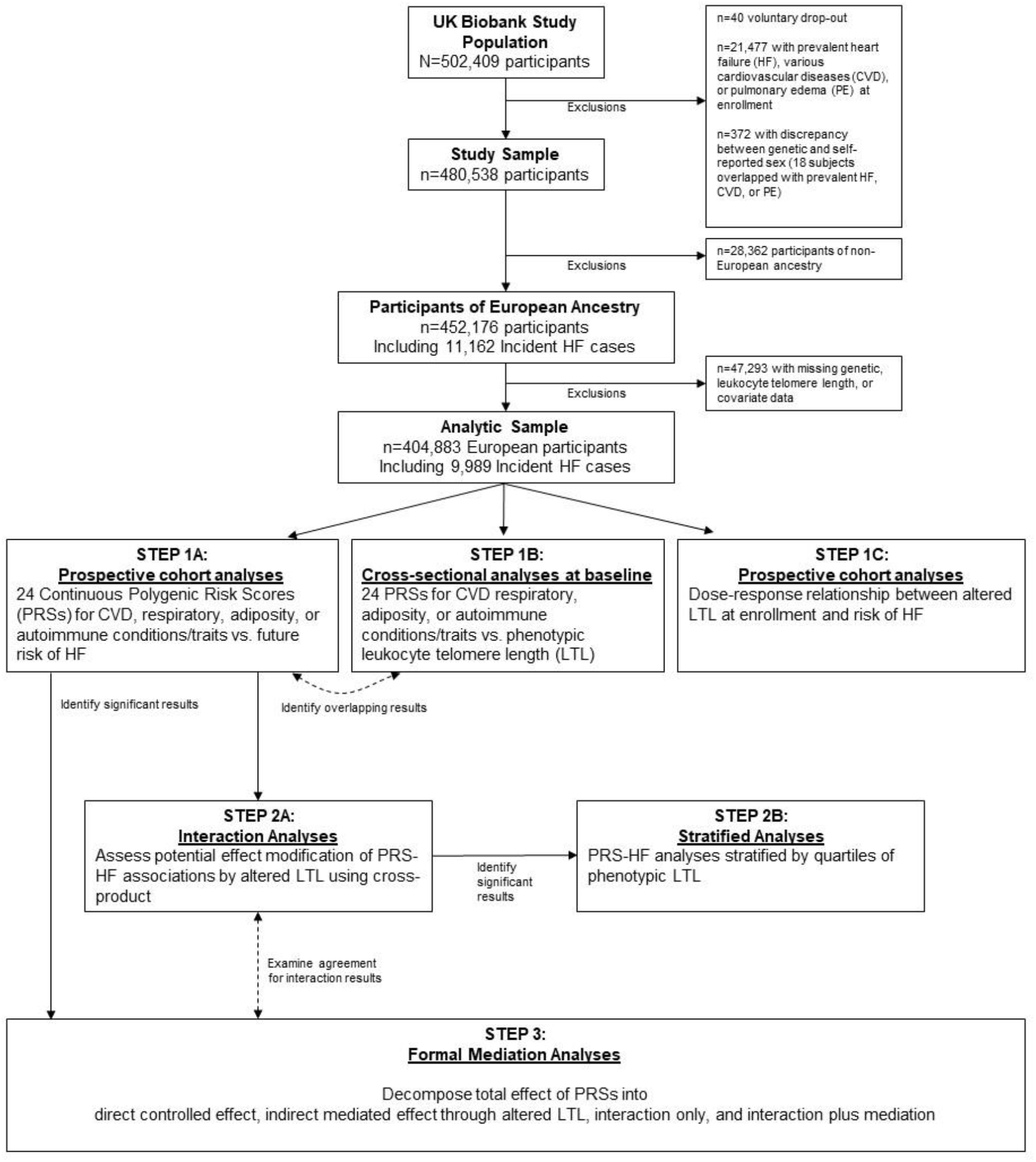
Multi-step analytical approach to investigate the interrelationships between genetic susceptibility, phenotypic leukocyte telomere length, and future risk of heart failure in the prospective UK Biobank.

## Results

### Dose-response relationships between polygenic risk scores for various diseases or traits and risk of heart failure

Among 404,883 participants of European ancestry, we identified 9,989 incident cases of HF (first reported in-patient hospitalization) throughout the 12.3 (1.8 SD) year follow-up. Among the HF cases, the average follow-up time to HF diagnosis was 8.3 (3.3 SD) years and the average age at diagnosis was 70.3 (7.0 SD) years.

First, we assessed the associations between 24 CVD-, cancer-, metabolic-, and inflammation-related polygenic risk scores (PRSs) ^42^ and future risk of developing HF (Figure 1) using multivariable Cox regression models adjusted for potential confounders as well as four genetic principal components to control for population stratification. The genome-wide data used to generate the PRSs were from external GWAS independent of the UK Biobank. We identified 9 PRSs that were significantly associated with HF risk that are reflective of increased genetic susceptibility to various cardiovascular diseases and traits, a respiratory disease, adiposity, and an autoimmune disease (Table 1). Notably, we found that increased genetic susceptibility to asthma, a prominent respiratory and inflammatory disease, was significantly associated with increased HF risk (P=1.8E-08; Table 1).

**Table 1:**
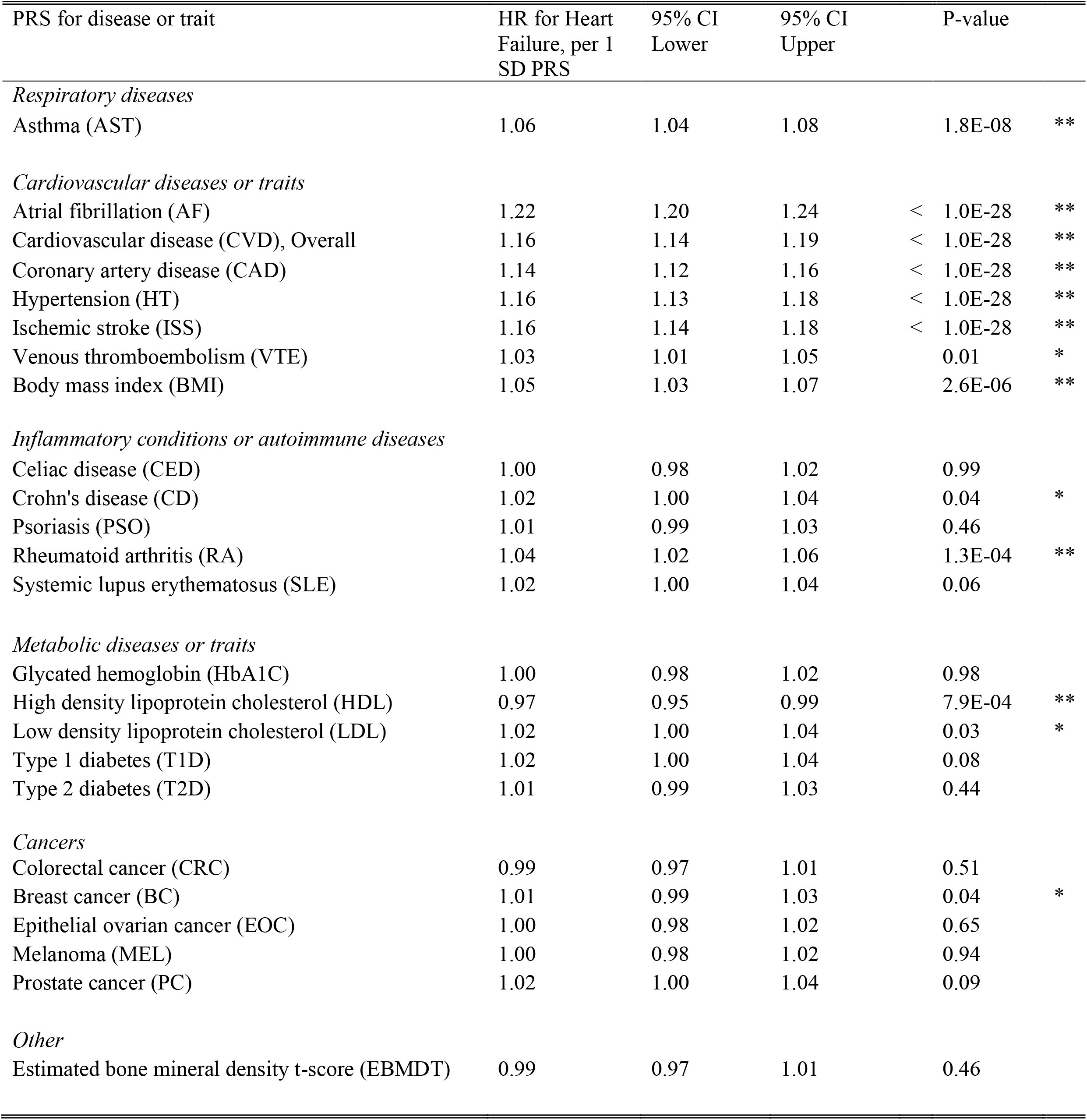
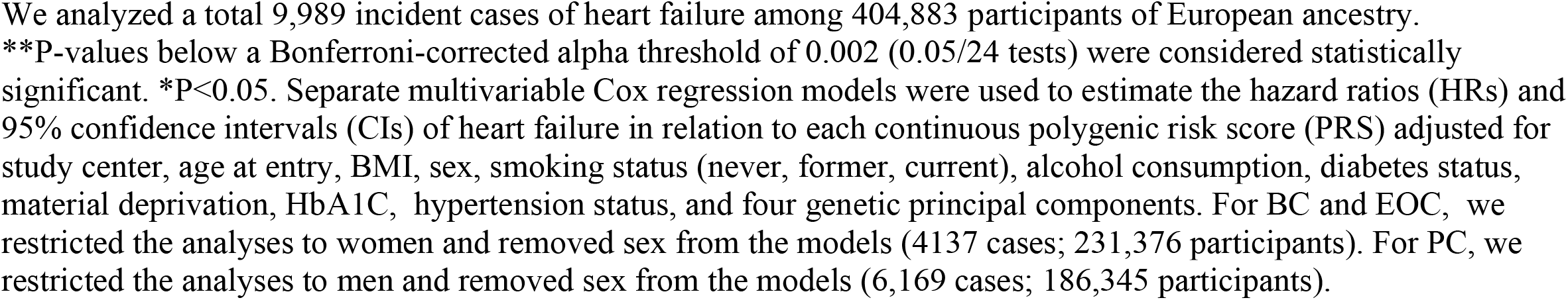
Polygenic risk scores for various diseases or traits and risk of heart failure among Europeans.

As expected, among the PRSs for cardiovascular diseases and traits, increased genetic susceptibility to AF (P<1.0E-28), overall CVD (P<1.0E-28), coronary artery disease (CAD; P<1.0E-28), hypertension (HT; P<1.0E-28), and ischemic stroke (ISS; P<1.0E-28) were significantly associated with elevated risk of HF in a dose-dependent manner (Table 1). However, the positive association between genetic susceptibility to venous thromboembolism (VTE) and HF risk was not significant after correction for multiple comparisons (P=0.01).

When examining autoimmune disorders, we found a positive dose-response relationship between genetic susceptibility to rheumatoid arthritis (RA) and HF risk (P=1.3E-04). However, associations with Crohn’s Disease (CD) were not significant after Bonferroni correction for multiple comparisons (P=0.04; Table 1). With respect to adiposity, genetically-predicted body mass index (BMI) was positively associated with HF risk (P=2.6E-06). When examining circulating cholesterol, increased genetically-predicted high-density lipoprotein (HDL) levels were significantly associated with lower risk of HF (P=7.9E-04), while increased low-density lipoprotein (LDL) levels were non-significantly associated with higher HF risk (P=0.03; Table 1). With respect to malignant diseases and HF risk, we did not detect any significant associations for genetic susceptibility to colorectal cancer (CRC), breast cancer (BC) and ovarian cancer (EOC) among women, and prostate cancer (PC) among men (Table 1).

### LTL modifies the effect of asthma genetic susceptibility on HF risk

Longer measured LTL, which reflects less exposure to environmental stressors, was significantly associated with decreased risk of HF (HR_per 1 SD LTL_=0.94, 95% CI: 0.92-0.96, P-trend=1.7E-08) independent of the PRSs. We then considered the 9 PRSs that were previously identified to be significantly associated with HF risk (Table 1) and tested for multiplicative interaction with measured LTL (Table 2). We found evidence for a super-multiplicative interaction between measured LTL and the PRS for asthma susceptibility, with longer LTL strengthening the positive asthma-HF association (P-interaction=2.8E-03). Additional Cox regression analyses stratified by quartiles of measured LTL supported a similar strengthening of the asthma-HF association with progressively longer LTL (Figure 2). However, we did not detect evidence for multiplicative interactions between LTL and the other PRSs (Table 2).

**Table 2:**
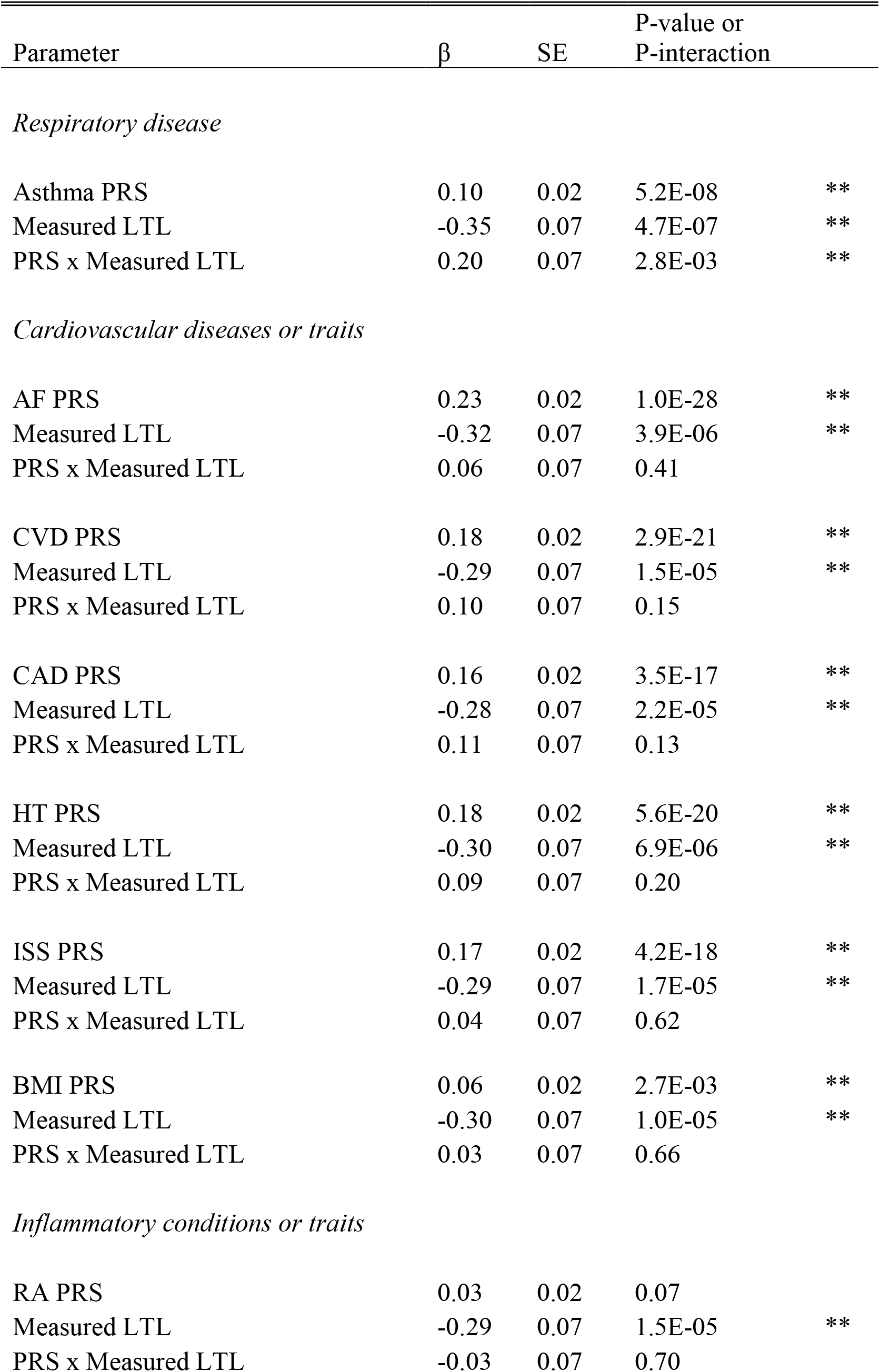
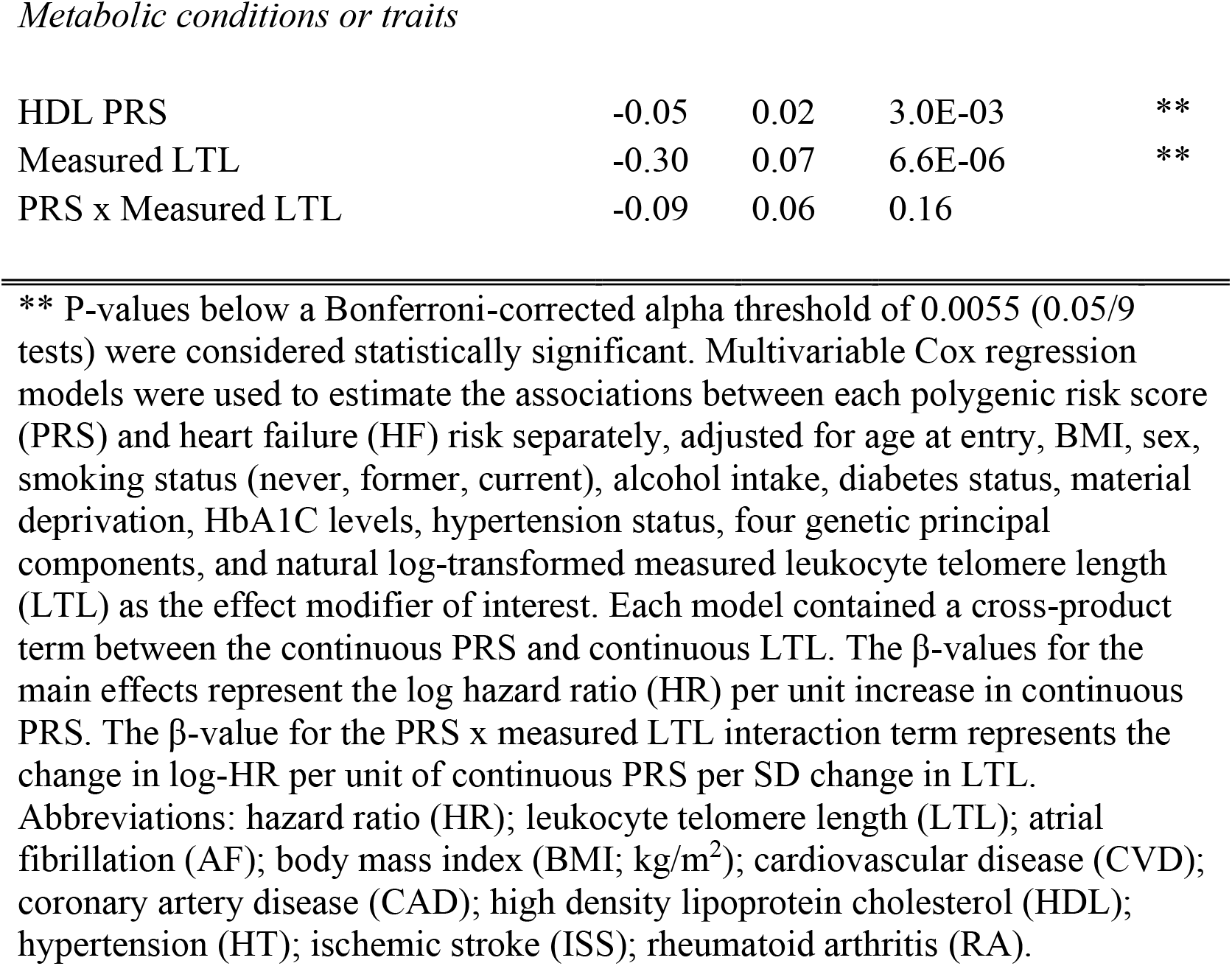
Multiplicative interaction between measured leukocyte telomere length and significant polygenic risk scores associated with HF risk.

| Parameter | $\beta$ | SE | P-value or<br>P-interaction | |
| --- | --- | --- | --- | --- |
| *Respiratory disease* |  |  |  |  |
| Asthma PRS | 0.10 | 0.02 | 5.2E-08 | ** |
| Measured LTL | -0.35 | 0.07 | 4.7E-07 | ** |
| PRS x Measured LTL | 0.20 | 0.07 | 2.8E-03 | ** |
| *Cardiovascular diseases or traits* |  |  |  |  |
| AF PRS | 0.23 | 0.02 | 1.0E-28 | ** |
| Measured LTL | -0.32 | 0.07 | 3.9E-06 | ** |
| PRS x Measured LTL | 0.06 | 0.07 | 0.41 |  |
| CVD PRS | 0.18 | 0.02 | 2.9E-21 | ** |
| Measured LTL | -0.29 | 0.07 | 1.5E-05 | ** |
| PRS x Measured LTL | 0.10 | 0.07 | 0.15 |  |
| CAD PRS | 0.16 | 0.02 | 3.5E-17 | ** |
| Measured LTL | -0.28 | 0.07 | 2.2E-05 | ** |
| PRS x Measured LTL | 0.11 | 0.07 | 0.13 |  |
| HT PRS | 0.18 | 0.02 | 5.6E-20 | ** |
| Measured LTL | -0.30 | 0.07 | 6.9E-06 | ** |
| PRS x Measured LTL | 0.09 | 0.07 | 0.20 |  |
| ISS PRS | 0.17 | 0.02 | 4.2E-18 | ** |
| Measured LTL | -0.29 | 0.07 | 1.7E-05 | ** |
| PRS x Measured LTL | 0.04 | 0.07 | 0.62 |  |
| BMI PRS | 0.06 | 0.02 | 2.7E-03 | ** |
| Measured LTL | -0.30 | 0.07 | 1.0E-05 | ** |
| PRS x Measured LTL | 0.03 | 0.07 | 0.66 |  |
| *Inflammatory conditions or traits* |  |  |  |  |
| RA PRS | 0.03 | 0.02 | 0.07 |  |
| Measured LTL | -0.29 | 0.07 | 1.5E-05 | ** |
| PRS x Measured LTL | -0.03 | 0.07 | 0.70 |  |
| *Metabolic conditions or traits* |  |  |  |  |
| HDL PRS | -0.05 | 0.02 | 3.0E-03 | ** |
| Measured LTL | -0.30 | 0.07 | 6.6E-06 | ** |
| PRS x Measured LTL | -0.09 | 0.06 | 0.16 |  |

\*\* P-values below a Bonferroni-corrected alpha threshold of 0.0055 (0.05/9 tests) were considered statistically significant. Multivariable Cox regression models were used to estimate the associations between each polygenic risk score (PRS) and heart failure (HF) risk separately, adjusted for age at entry, BMI, sex, smoking status (never, former, current), alcohol intake, diabetes status, material deprivation, HbA1C levels, hypertension status, four genetic principal components, and natural log-transformed measured leukocyte telomere length (LTL) as the effect modifier of interest. Each model contained a cross-product term between the continuous PRS and continuous LTL. The $\beta$ -values for the main effects represent the log hazard ratio (HR) per unit increase in continuous PRS. The $\beta$ -value for the PRS x measured LTL interaction term represents the change in log-HR per unit of continuous PRS per SD change in LTL. Abbreviations: hazard ratio (HR); leukocyte telomere length (LTL); atrial fibrillation (AF); body mass index (BMI; kg/m<sup>2</sup>); cardiovascular disease (CVD); coronary artery disease (CAD); high density lipoprotein cholesterol (HDL); hypertension (HT); ischemic stroke (ISS); rheumatoid arthritis (RA).

**Figure 2:**
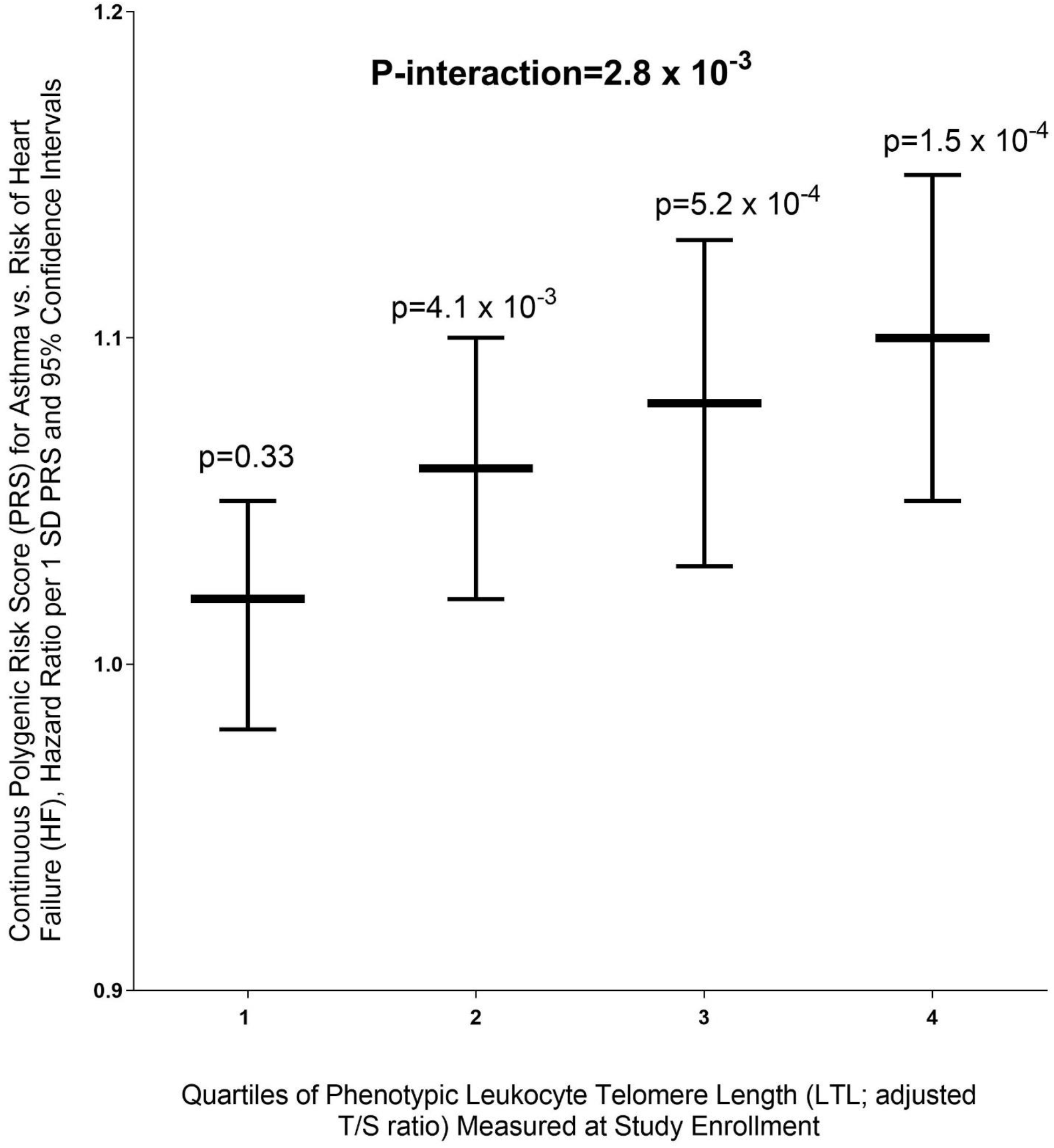
Phenotypic leukocyte telomere length measured at enrollment interacts with genetic susceptibility to asthma to strengthen its association with heart failure risk in the UK Biobank.

### The effect of genetic susceptibility to various disease or traits on LTL measured at enrollment

We assessed whether the 24 PRSs were associated with altered LTL measured at enrollment and identified 13 significant findings (Table 3). We found that increased genetic susceptibility to asthma (P=1.7E-06), overall CVD (P=3.0E-18), celiac disease (CED; P<1.0E-28), CAD (P= 6.5E-13), ISS (P=1.2E-06), systemic lupus erythematosus (SLE; P= 7.2E-15), type 1 diabetes (T1D; P= 8.8E-10), and type 2 diabetes (T2D; P= 1.8E-06) as well as genetically predicted HbA1C levels (P= 1.2E-04) were associated with shorter LTL, which reflects increased biological aging (Table 3). Additionally, we found that higher genetic susceptibility to EOC (P= 3.8E-15), melanoma (MEL; P< 1.0E-28), and PC (P= 6.8E-13) as well as genetically-predicted LDL levels (6.2E-06) were associated with longer LTL (Table 3). Taken together, these findings suggest shared genetic etiology between these diseases or traits with the biological aging component of LTL (Table 3). Among these 13 significant findings, the PRSs for asthma, CVD, CAD, and ISS overlapped with the significant associations identified in the PRS-HF analyses (Table 1), which suggests potential interrelationships between genetic susceptibility to cardiovascular and pulmonary outcomes, phenotypic LTL, and future risk of HF. These findings prompted us to assess whether the effects of the 9 previously identified PRSs on HF risk (Table 1) were potentially mediated through altered LTL (Figure 1).

**Table 3:**
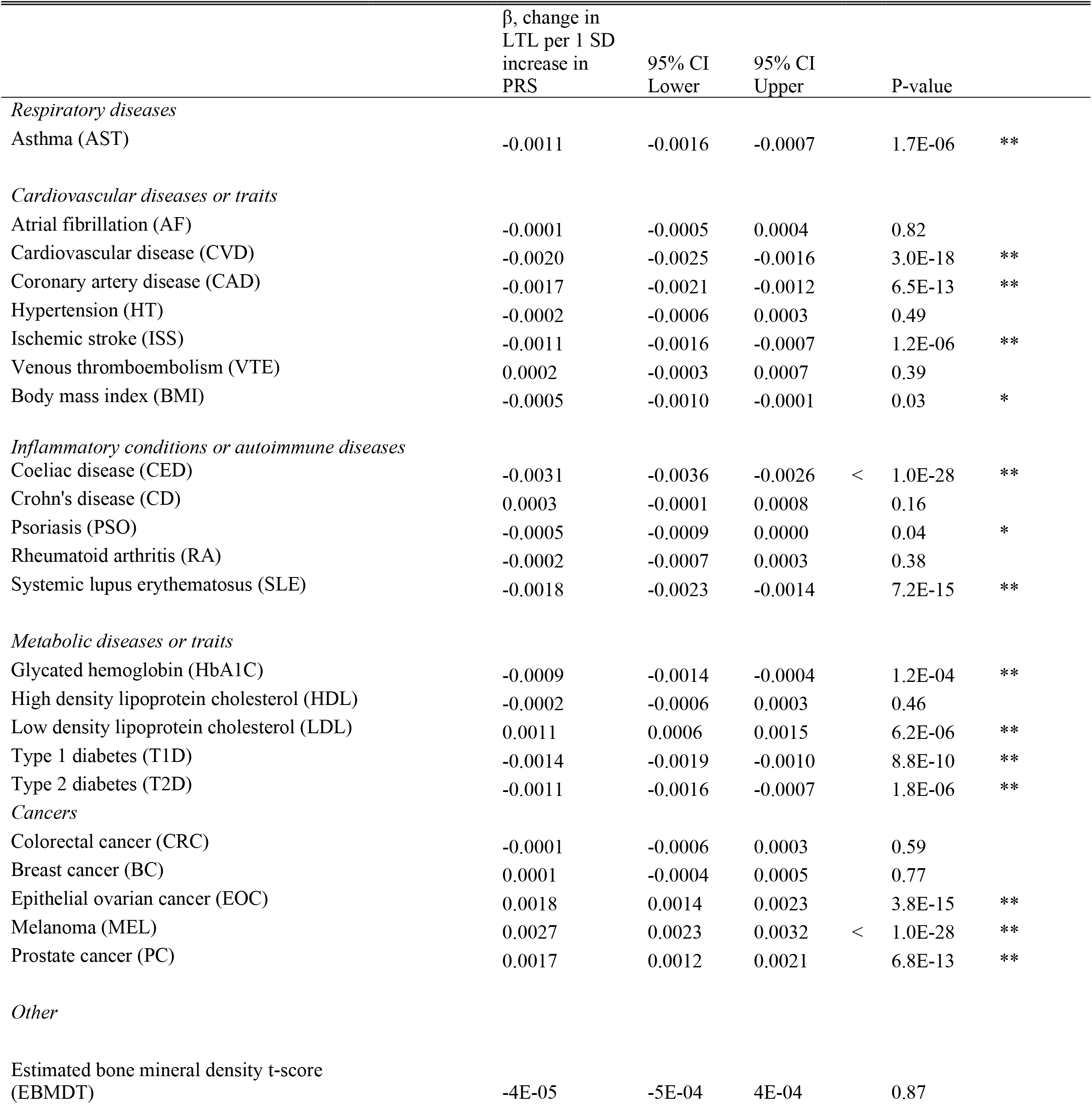
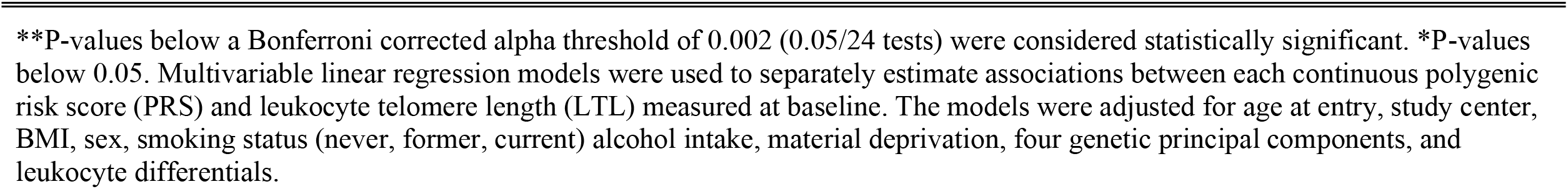
Associations between polygenic risk scores for various diseases and measured leukocyte telomere length measured at enrollment.

### Limited evidence for LTL as a mediator of PRS-HF associations

We conducted formal mediation analyses, which decomposes the total effect of each of the PRSs on HF risk into four components: 1) the pure direct effect; 2) only interaction, but not mediation; 3) both mediation and interaction; and 4) only mediation, but not interaction (i.e., pure indirect effect) ^43^. To facilitate the mediation analyses, we dichotomized each of the 9 PRSs associated with HF risk by their respective 75^th^ percentiles into a “high” genetic susceptibility category and a “normal-to-low” genetic susceptibility category (referent). Among the 9 PRSs, we found 8 that were associated with HF with statistically significant total excess relative risks (Table 4). The 8 noteworthy PRSs included those related to asthma, AF, BMI, CVD, CAD, HDL, HT, and ISS. Among these 8 PRSs, we were primarily interested in the excess relative risk due to the pure indirect effect through LTL (i.e., the mediated effect). The strongest mediated effect through LTL was for the asthma PRS, which accounted for a very small but statistically significant 1.13% of the total effect (P<0.001; Table 4). The mediated effect through LTL for the BMI, CVD, CAD, HT, and ISS PRSs were also statistically significant (P<0.001) but accounted for only 0.23% - 0.60% of their respective total effects (Table 4).

**Table 4:**
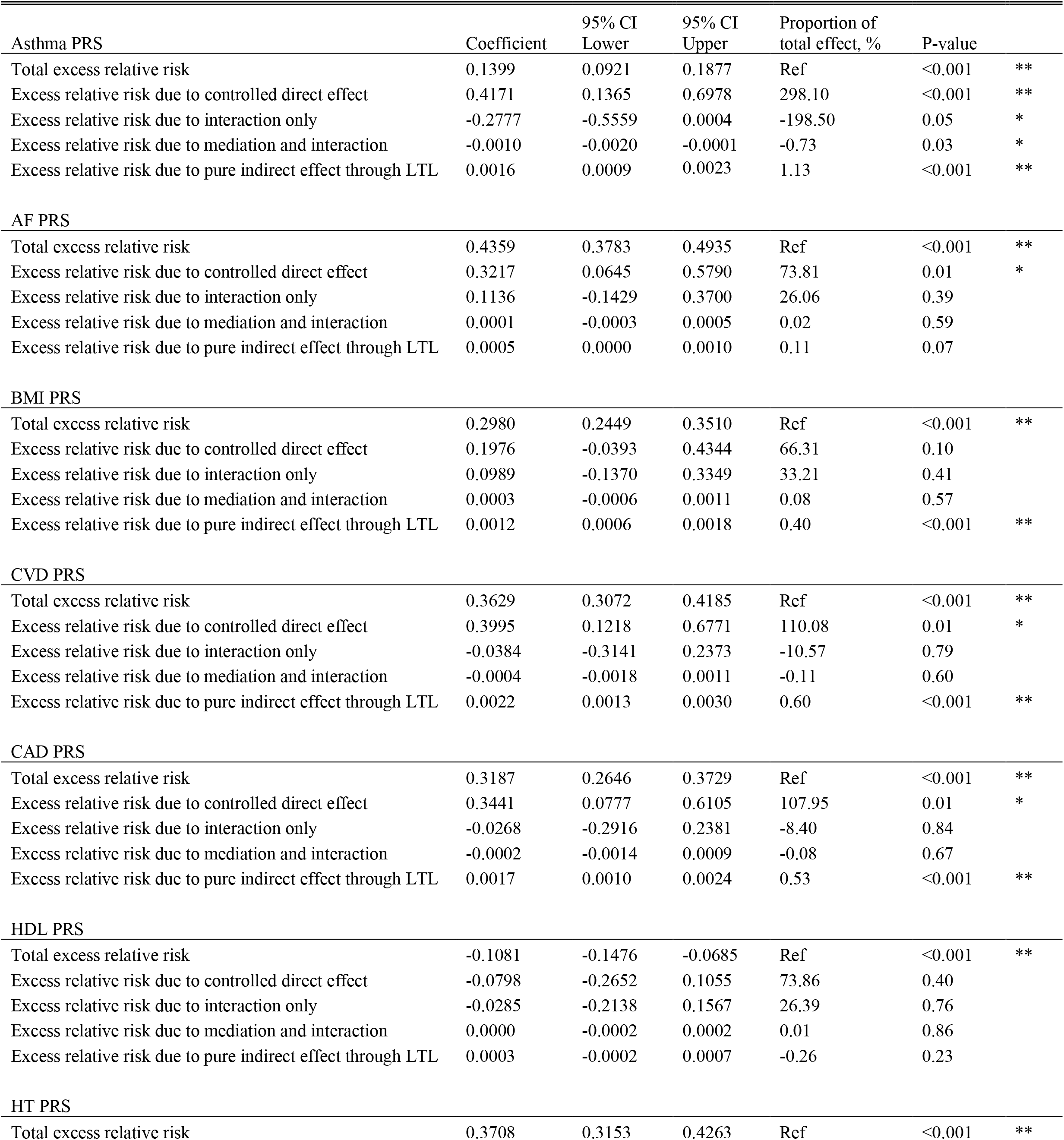
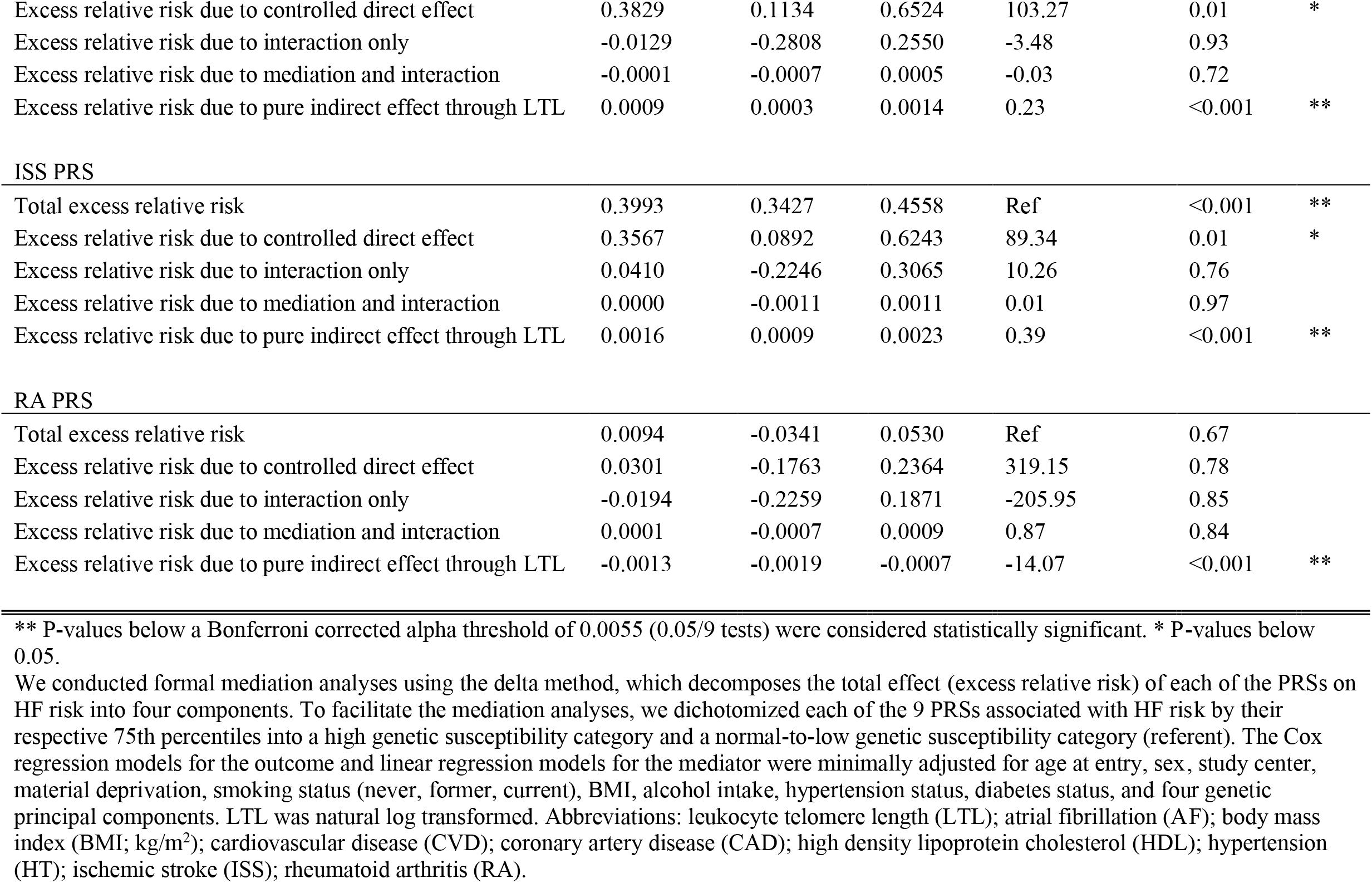
Mediation analyses to decompose the total effect of polygenic risk scores on heart failure risk through altered leukocyte telomere length.

|  | Coefficient | 95% CI<br>Lower | 95% CI<br>Upper | Proportion of<br>total effect, % | P-value |  |
| --- | --- | --- | --- | --- | --- | --- |
| **Asthma PRS** |  |  |  |  |  |  |
| Total excess relative risk | 0.1399 | 0.0921 | 0.1877 | Ref | <0.001 | ** |
| Excess relative risk due to controlled direct effect | 0.4171 | 0.1365 | 0.6978 | 298.10 | <0.001 | ** |
| Excess relative risk due to interaction only | -0.2777 | -0.5559 | 0.0004 | -198.50 | 0.05 | * |
| Excess relative risk due to mediation and interaction | -0.0010 | -0.0020 | -0.0001 | -0.73 | 0.03 | * |
| Excess relative risk due to pure indirect effect through LTL | 0.0016 | 0.0009 | 0.0023 | 1.13 | <0.001 | ** |
| **AF PRS** |  |  |  |  |  |  |
| Total excess relative risk | 0.4359 | 0.3783 | 0.4935 | Ref | <0.001 | ** |
| Excess relative risk due to controlled direct effect | 0.3217 | 0.0645 | 0.5790 | 73.81 | 0.01 | * |
| Excess relative risk due to interaction only | 0.1136 | -0.1429 | 0.3700 | 26.06 | 0.39 |  |
| Excess relative risk due to mediation and interaction | 0.0001 | -0.0003 | 0.0005 | 0.02 | 0.59 |  |
| Excess relative risk due to pure indirect effect through LTL | 0.0005 | 0.0000 | 0.0010 | 0.11 | 0.07 |  |
| **BMI PRS** |  |  |  |  |  |  |
| Total excess relative risk | 0.2980 | 0.2449 | 0.3510 | Ref | <0.001 | ** |
| Excess relative risk due to controlled direct effect | 0.1976 | -0.0393 | 0.4344 | 66.31 | 0.10 |  |
| Excess relative risk due to interaction only | 0.0989 | -0.1370 | 0.3349 | 33.21 | 0.41 |  |
| Excess relative risk due to mediation and interaction | 0.0003 | -0.0006 | 0.0011 | 0.08 | 0.57 |  |
| Excess relative risk due to pure indirect effect through LTL | 0.0012 | 0.0006 | 0.0018 | 0.40 | <0.001 | ** |
| **CVD PRS** |  |  |  |  |  |  |
| Total excess relative risk | 0.3629 | 0.3072 | 0.4185 | Ref | <0.001 | ** |
| Excess relative risk due to controlled direct effect | 0.3995 | 0.1218 | 0.6771 | 110.08 | 0.01 | * |
| Excess relative risk due to interaction only | -0.0384 | -0.3141 | 0.2373 | -10.57 | 0.79 |  |
| Excess relative risk due to mediation and interaction | -0.0004 | -0.0018 | 0.0011 | -0.11 | 0.60 |  |
| Excess relative risk due to pure indirect effect through LTL | 0.0022 | 0.0013 | 0.0030 | 0.60 | <0.001 | ** |
| **CAD PRS** |  |  |  |  |  |  |
| Total excess relative risk | 0.3187 | 0.2646 | 0.3729 | Ref | <0.001 | ** |
| Excess relative risk due to controlled direct effect | 0.3441 | 0.0777 | 0.6105 | 107.95 | 0.01 | * |
| Excess relative risk due to interaction only | -0.0268 | -0.2916 | 0.2381 | -8.40 | 0.84 |  |
| Excess relative risk due to mediation and interaction | -0.0002 | -0.0014 | 0.0009 | -0.08 | 0.67 |  |
| Excess relative risk due to pure indirect effect through LTL | 0.0017 | 0.0010 | 0.0024 | 0.53 | <0.001 | ** |
| **HDL PRS** |  |  |  |  |  |  |
| Total excess relative risk | -0.1081 | -0.1476 | -0.0685 | Ref | <0.001 | ** |
| Excess relative risk due to controlled direct effect | -0.0798 | -0.2652 | 0.1055 | 73.86 | 0.40 |  |
| Excess relative risk due to interaction only | -0.0285 | -0.2138 | 0.1567 | 26.39 | 0.76 |  |
| Excess relative risk due to mediation and interaction | 0.0000 | -0.0002 | 0.0002 | 0.01 | 0.86 |  |
| Excess relative risk due to pure indirect effect through LTL | 0.0003 | -0.0002 | 0.0007 | -0.26 | 0.23 |  |
| **HT PRS** |  |  |  |  |  |  |
| Total excess relative risk | 0.3708 | 0.3153 | 0.4263 | Ref | <0.001 | ** |
| Excess relative risk due to controlled direct effect | 0.3829 | 0.1134 | 0.6524 | 103.27 | 0.01 | * |
| Excess relative risk due to interaction only | -0.0129 | -0.2808 | 0.2550 | -3.48 | 0.93 |  |
| Excess relative risk due to mediation and interaction | -0.0001 | -0.0007 | 0.0005 | -0.03 | 0.72 |  |
| Excess relative risk due to pure indirect effect through LTL | 0.0009 | 0.0003 | 0.0014 | 0.23 | <0.001 | ** |
| ISS PRS |  |  |  |  |  |  |
| Total excess relative risk | 0.3993 | 0.3427 | 0.4558 | Ref | <0.001 | ** |
| Excess relative risk due to controlled direct effect | 0.3567 | 0.0892 | 0.6243 | 89.34 | 0.01 | * |
| Excess relative risk due to interaction only | 0.0410 | -0.2246 | 0.3065 | 10.26 | 0.76 |  |
| Excess relative risk due to mediation and interaction | 0.0000 | -0.0011 | 0.0011 | 0.01 | 0.97 |  |
| Excess relative risk due to pure indirect effect through LTL | 0.0016 | 0.0009 | 0.0023 | 0.39 | <0.001 | ** |
| RA PRS |  |  |  |  |  |  |
| Total excess relative risk | 0.0094 | -0.0341 | 0.0530 | Ref | 0.67 |  |
| Excess relative risk due to controlled direct effect | 0.0301 | -0.1763 | 0.2364 | 319.15 | 0.78 |  |
| Excess relative risk due to interaction only | -0.0194 | -0.2259 | 0.1871 | -205.95 | 0.85 |  |
| Excess relative risk due to mediation and interaction | 0.0001 | -0.0007 | 0.0009 | 0.87 | 0.84 |  |
| Excess relative risk due to pure indirect effect through LTL | -0.0013 | -0.0019 | -0.0007 | -14.07 | <0.001 | ** |

\*\* P-values below a Bonferroni corrected alpha threshold of 0.0055 (0.05/9 tests) were considered statistically significant. \* P-values below 0.05.
We conducted formal mediation analyses using the delta method, which decomposes the total effect (excess relative risk) of each of the PRSs on HF risk into four components. To facilitate the mediation analyses, we dichotomized each of the 9 PRSs associated with HF risk by their respective 75th percentiles into a high genetic susceptibility category and a normal-to-low genetic susceptibility category (referent). The Cox regression models for the outcome and linear regression models for the mediator were minimally adjusted for age at entry, sex, study center, material deprivation, smoking status (never, former, current), BMI, alcohol intake, hypertension status, diabetes status, and four genetic principal components. LTL was natural log transformed. Abbreviations: leukocyte telomere length (LTL); atrial fibrillation (AF); body mass index (BMI; kg/m<sup>2</sup>); cardiovascular disease (CVD); coronary artery disease (CAD); high density lipoprotein cholesterol (HDL); hypertension (HT); ischemic stroke (ISS); rheumatoid arthritis (RA).

## Discussion

### Shared genetic etiology between various chronic diseases or traits and HF risk

We conducted comprehensive analyses to investigate the shared genetic etiology between various chronic diseases or traits and HF risk in a large-scale prospective cohort study of Europeans. First, we found that increased genetic susceptibility to cardiovascular, respiratory, autoimmune, and adiposity related diseases or traits was associated with future risk of HF in a population without clinically diagnosed CVD at enrollment. These findings suggest shared genetic etiology between these diseases or traits and the pathogenesis of HF. The positive findings for CVD are consistent with the fact that HF is a terminal manifestation of most forms of CVD. The findings for RA were of particular interest because of the growing appreciation of inflammatory or autoimmune disorders in HF pathogenesis ^44^, while asthma was of interest because of the established link between pulmonary diseases, cardiac function, and inflammation.

### The interrelationships between asthma genetic susceptibility, LTL, and HF risk

To gain additional mechanistic insight, we also investigated how LTL influences the effect of genetic susceptibility on HF risk. In agreement with previous analyses ^18, 31^, we found that longer phenotypic LTL, which reflects decreased environmental stressors and biological aging, was associated with lower risk of HF. Subsequently, we found evidence that longer phenotypic LTL acted as an effect modifier that strengthened the positive association between asthma genetic susceptibility and HF risk. This effect modification was likely attributed to the environmentally-determined components reflected by LTL, which explain most of the variance in telomere length. These findings support the potential utility of incorporating LTL and genetic data into future risk stratification analyses. However, we found negligible evidence for indirect effects mediated through the biological components reflected by LTL. As such, the environmental components reflected by LTL may be of greater etiologic importance to HF compared to the biological or genetic components.

### The genetic link between asthma and HF risk

Asthma is a chronic respiratory and inflammatory disease that is characterized by obstructed airflow and bronchospasms ^45^. Previous studies have found some evidence linking asthma to HF ^46^ and its risk factors ^47^. A cohort study conducted in Northern California found that those with asthma had over double the risk of HF ^48^. Additionally, a Mendelian Randomization (MR) analysis conducted in a European population from FinnGen and the Heart Failure Molecular Epidemiology for Therapeutic targets consortium found evidence suggesting a causal association between asthma and HF risk ^49^. Our PRS findings confirm the previously conducted MR analyses ^49^ and expand upon the molecular mechanism by elucidating the role of LTL as an effect modifier.

Even though phenotypic LTL was inversely associated with HF independent of genetic susceptibility, it had a super-multiplicative interaction with the asthma PRS to strengthen the association with HF. The reason for the direction of this statistical interaction is unclear. Given that phenotypic LTL reflects endogenous biological processes related to aging as well as cumulative burden of exogenous environmental exposures, it is possible that each genetic variant used to derive the PRS could be interacting differently with the various components reflected by LTL. Complementary functional experiments are needed to further elucidate the nature of any potential biological interaction.

### Genetic liability to other non-malignant and malignant diseases and their role in altering LTL

In addition to the main results, we also found that genetic susceptibility to various cardiovascular diseases and traits as well as cancers was associated with altered phenotypic LTL measured at the time of enrollment. The observed relationships between cardiovascular disease and traits and *shorter* LTL were consistent with previous prospective and retrospective studies ^33, 50^ and further support the role of their shared genetics ^51, 52^. Interestingly, we also found that genetic susceptibility to melanoma, ovarian epithelial cancer, and prostate cancer was associated with *longer* phenotypic LTL, which was in concordance with previous MR analyses in European and Asian populations ^34^. However, the relationship between telomere length and cancer risk is complex and nuanced. In prospective studies, LTL associations were equivocal for risk of breast cancer and other sites, ^35, 37, 53, 54^; *shorter* LTL was associated with increased risk of bladder ^55^, esophageal ^56^ and overall cancer ^57^; and *longer* LTL was associated with increased risk of non-Hodgkin lymphoma ^58^ and potentially melanoma ^59^. Most notably, *longer* phenotypic and genetically-determined LTL has been consistently found to be associated with *increased* risk of lung adenocarcinoma among Europeans and East Asians ^34, 38–40, 60–63^. These positive associations defy the traditional expectation of telomere dynamics in cancer development and the precise biological mechanism underlying these paradoxical relationships is unclear. A potential explanation is that longer telomeres in target tissue (indirectly reflected by LTL) may lead to delayed senescence of pre-cancerous cells, which facilitates the accumulation of somatic abnormalities that potentially drive carcinogenesis and cancer progression ^40, 60 20^.

### Strengths and limitations

Our study had numerous strengths. First, the UK Biobank is the world’s largest prospective cohort study to date that measured LTL in pre-diagnostic biospecimens. The sample size and rich data optimized the statistical power to detect complex associations, which is particularly important when investigating statistical interactions. Furthermore, the prospective study design allowed temporality to be established between genetic susceptibility, phenotypic LTL, and HF risk, which permitted mediation analyses to evaluate the precise interrelationships between these variables. Additionally, the PRSs used in our analyses were derived for the UK Biobank using a Bayesian method with genetic variants across the entire genome, as opposed to conventional PRSs which are comprised of a limited number of genome-wide significant variants ^42^. Lastly, the UK Biobank was linked to national hospital registries, which likely captured most HF cases in the cohort, thus reducing the potential for outcome misclassification.

Our study had some limitations. Most of the UK Biobank participants were of White European ancestry, which limited the generalizability of our findings. The observed interrelationships could be potentially different among ethnic subgroups with different genomic architecture and exposure profiles that influence LTL and HF risk. Additionally, we had limited information on heart failure subtypes, severity, and whether left ventricle ejection fraction was preserved or not. Lastly, phenotypic LTL measured using qPCR has become a mainstay among epidemiologic studies of chronic disease. However, the adjusted relative telomere-to-single copy gene (T/S ratio) reflects the average telomere length in a population of white blood cells but does not provide information on the absolute telomere length of specific chromosomal arms.

In summary, our findings strongly suggest that people with both longer LTL and increased genetic liability to asthma may have elevated risk of HF, a highly fatal consequence of cardiac dysfunction that has a mortality rate comparable to some cancers ^64 65^. These findings contribute to the understanding of the shared molecular/genetic mechanisms underlying HF and non-malignant respiratory diseases, as well as the role of LTL as an effect modifier in the underlying mechanisms. Importantly, we potentially identified higher-risk subpopulations for HF, which warrants further investigation of LTL and genetics in future risk stratification analyses.

## Data Availability

Data used for this study are publicly available at: https://bbams.ndph.ox.ac.uk/ams/ Meta-data and codes produced in the present study are available upon reasonable request to the authors.

## Acknowledgements

This study was supported by intramural funding from the National Cancer Institute and National Heart, Lung, and Blood Institute. We thank Drs. Qing Lan and Nathaniel Rothman for their continuing support. Additionally, we extend our deepest appreciation to Lisa Finkelstein, Jillian Varonin, and the UK Biobank Access Team for helping us navigate complex international data agreements. We declare no conflicts of interest.

## CRediT author statement

Jason Y.Y. Wong-Conceptualization, methodology, formal analysis, data curation, writing original draft

Batel Blechter-Methodology, formal analysis, writing review and editing

Zhonghua Liu-Methodology, writing review and editing

Jianxin Shi-Methodology, formal analysis, writing review and editing

Véronique L. Roger-Supervision, conceptualization, writing original draft

## Online Supplement

### Methods

#### Study design

The UK Biobank is a prospective cohort study with extensive biospecimen collection at the time of enrollment. The study design, rationale, and methodologies have been described in detail ^66, 67^. In brief, the source population was adults aged 40-69 years who lived ≤40 km of 22 study assessment centers throughout the UK. Approximately 9.2 million people registered in the UK’s National Health Service were mailed study invitations, whereas a subset of 503,317 people (5.5%) visited the assessment centers during the recruitment period of 2006-2010 ^66^. Volunteer participants who provided electronic informed consent were given touchscreen questionnaires, physical examinations, and provided biospecimens for biomarker analyses. As part of UK Biobank project number: 28072, our dataset obtained in August 2022 included a total of 502,409 participants. The UK Biobank is a continuously updated dataset without periodic data freezes.

#### Heart failure diagnosis

Heart failure was defined using in-patient hospital diagnoses coded according to the International Classification of Disease version 9 (ICD-9) and 10 (ICD-10) classifications. ICD-10 codes I50.0, I50.1, I50.9, I11.0, I13.0, and I13.2 along with ICD-9 codes 428.0 and 428.1 were used to define heart failure.

#### Prospective follow-up

For each participant, follow-up time started at the date of visit to the assessment center in 2006-2010 and ended at the date of first incident heart failure diagnosis (outcome), death (censored), or administrative censoring (i.e., September 20^th^, 2021, for England and Wales and October 31^st^, 2021, for Scotland), whichever came first. Vital status as well as date and primary underlying cause of death was provided by the NHS Information Centre and the NHS Central Register Scotland. The UK Biobank study was approved by the National Information Governance Board for Health and Social Care and the NHS North West Multicenter Research Ethics Committee. All volunteer participants provided electronic informed consent.

#### Measured phenotypic leukocyte telomere length (LTL)

DNA extraction, LTL measurement, and quality control were performed as previously described ^68^. Data from the multiplex quantitative polymerase chain reaction (qPCR) assay were used to calculate the ratio of telomere repeat copy number (T) to single-copy gene copy number (S; beta-globin) for each subject. This ‘T/S ratio’ for each subject was then divided by the ‘T/S ratio’ of a calibrator sample included on every run to calculate the ‘*relative* T/S ratio’, which reflects the average telomere abundance across all chromosomes in leukocytes of an individual. The ‘*adjusted relative* T/S ratio’ accounted for batch variation over time ^68^.

#### Polygenic Risk Scores (PRSs) for various diseases and quantitative traits

The 24 PRSs (a subset of 28 diseases and 8 quantitative traits) used in our analyses were previously derived as part of UK Biobank project number 9659 as described in detail (https://biobank.ndph.ox.ac.uk/ukb/label.cgi?id=301) ^42^. Briefly, genotype data from 825,927 genetic variants were generated using a custom Axiom genotyping array, followed by genome-wide imputation. The PRS algorithms were constructed from trait-specific meta-analyses using a Bayesian approach combining data across multiple ancestries and related traits where appropriate ^42^. The PRS values for each participant were calculated as the genome-wide sum of the per-variant posterior effect size multiplied by allele dosage ^42^. A principal component (PC)-based ancestry centering step ^69^ was used to center the PRS distributions across all ancestries. PRS distributions were also standardized to have approximately unit variance within ancestry groups.

The Bayesian PRSs were generated by Genomics plc (https://www.genomicsplc.com/) in partnership with Our Future Health (https://ourfuturehealth.org.uk/). The performance metrics and validation of these PRSs were as described elsewhere ^42^. Additional meta data, datasets, and code can be found at: https://github.com/Genomicsplc/ukb-pret/blob/main/UK_Biobank_PRS_Release_Dataset_Notes.md, https://biobank.ndph.ox.ac.uk/showcase/refer.cgi?id=5202, and https://zenodo.org/record/6631952. The genetic data used to derive the standard PRSs were from external GWAS data independent of the UK Biobank. The standard PRSs were generated for all UK Biobank participants. The ‘enhanced’ PRSs previously derived by Thompson et al. ^42^, which incorporates UK Biobank and external data, were not analyzed in our study because they were only available for a subset of the participants and undergoing further development.

#### Statistical analyses

##### Associations between PRSs and HF risk

Multivariable Cox regression models were used to estimate the hazard ratios (HRs) and 95% confidence intervals (CIs) of incident diagnosed heart failure (first reported in-patient hospital admission), in relation to the per 1 SD increase of each of the 24 continuous PRSs in separate analyses. The models were further adjusted for potential confounders including study assessment center, age at entry, body mass index (BMI), sex, smoking status (never, former, current), alcohol consumption, diabetes status, material deprivation (continuous Townsend deprivation index), HbA1C levels, hypertension status, and four genetic principal components. For breast cancer and epithelial ovarian cancer PRSs, we restricted the analyses to women and removed sex from the models (4137 cases; 231,376 participants). For the prostate cancer PRS, we restricted the analyses to men and removed sex from the models (6,169 cases; 186,345 participants). Follow-up time was used as the underlying timescale to improve consistency with previous analyses ^70, 71^. Evidence for multiplicative effect modification was assessed using cross-product terms in the models between the PRSs and continuous natural log-transformed LTL. We conducted additional PRS-HF analyses stratified by quartiles of LTL. Proportional hazards assumptions were evaluated using Supremum tests and visually examining plots of Schoenfeld residuals over follow-up time. Bonferroni correction of the alpha significance threshold (i.e., 0.05 / number of tests) was used to account for multiple comparisons.

##### Associations between PRSs and LTL

Multivariable linear regression models were used to separately estimate associations between each continuous PRS and continuous natural log-transformed LTL measured at enrollment. The models were adjusted for age at entry, study center, BMI, sex, smoking status (never, former, current) alcohol intake, material deprivation, four genetic principal components, and leukocyte differentials.

##### Mediation analyses

We evaluated whether the association between the noteworthy PRSs and HF risk was mediated through continuous LTL using the Med4Way package in STATA/MP 13.0 (StataCorp LP, College Station, TX) ^43^. This approach uses the delta method to decompose the total effect of exposure on outcome into four components that correspond to the portion of the total effect that is due to: 1) pure direct effect of exposure on outcome; 2) interaction but not mediation; 3) both mediation and interaction ; and 4) pure indirect mediated effect ^43^. To facilitate the mediation analyses, we dichotomized each of the noteworthy PRSs by their respective 75th percentiles into a ‘high’ genetic susceptibility category and a ‘normal-to-low’ genetic susceptibility category (referent). The Cox regression models for the outcome and linear regression models for the mediator were minimally adjusted for age at entry, sex, study assessment center, material deprivation, smoking status (never, former, current), BMI, alcohol intake, hypertension status, diabetes status, and four genetic principal components. LTL was natural log-transformed to approximate a normal distribution.

## References

1. Jessup M, Abraham WT, Casey DE, Feldman AM, Francis GS, Ganiats TG, Konstam MA, Mancini DM, Rahko PS, Silver MA, et al. 2009 focused update: ACCF/AHA Guidelines for the Diagnosis and Management of Heart Failure in Adults: a report of the American College of Cardiology Foundation/American Heart Association Task Force on Practice Guidelines: developed in collaboration with the International Society for Heart and Lung Transplantation. Circulation. 2009;119:1977–2016. doi: 10.1161/CIRCULATIONAHA.109.192064

2. Swedberg K, Cleland J, Dargie H, Drexler H, Follath F, Komajda M, Tavazzi L, Smiseth OA, Gavazzi A, Haverich A, et al. Guidelines for the diagnosis and treatment of chronic heart failure: executive summary (update 2005): The Task Force for the Diagnosis and Treatment of Chronic Heart Failure of the European Society of Cardiology. Eur Heart J. 2005;26:1115–1140. doi: 10.1093/eurheartj/ehi204

3. Roger VL. Epidemiology of heart failure. Circ Res. 2013;113:646–659. doi: 10.1161/CIRCRESAHA.113.300268

4. Groenewegen A, Rutten FH, Mosterd A, Hoes AW. Epidemiology of heart failure. Eur J Heart Fail. 2020;22:1342–1356. doi: 10.1002/ejhf.1858

5. Roger VL. Epidemiology of Heart Failure: A Contemporary Perspective. Circ Res. 2021;128:1421–1434. doi: 10.1161/CIRCRESAHA.121.318172

6. Shah S, Henry A, Roselli C, Lin H, Sveinbjornsson G, Fatemifar G, Hedman AK, Wilk JB, Morley MP, Chaffin MD, et al. Genome-wide association and Mendelian randomisation analysis provide insights into the pathogenesis of heart failure. Nat Commun. 2020;11:163. doi: 10.1038/s41467-019-13690-5

7. Lindgren MP, PirouziFard M, Smith JG, Sundquist J, Sundquist K, Zoller B. A Swedish Nationwide Adoption Study of the Heritability of Heart Failure. JAMA Cardiol. 2018;3:703–710. doi: 10.1001/jamacardio.2018.1919

8. Lumbers RT, Shah S, Lin H, Czuba T, Henry A, Swerdlow DI, Malarstig A, Andersson C, Verweij N, Holmes MV, et al. The genomics of heart failure: design and rationale of the HERMES consortium. ESC Heart Fail. 2021;8:5531–5541. doi: 10.1002/ehf2.13517

9. Hamo CE, Bloom MW. Cancer and Heart Failure: Understanding the Intersection. Card Fail Rev. 2017;3:66–70. doi: 10.15420/cfr.2016:24:2

10. Fradley MG. Heart Failure in Patients With Cancer Treated With Anthracyclines-Revisiting the Foundation of Cardio-Oncology. JAMA Netw Open. 2023;6:e2254677. doi: 10.1001/jamanetworkopen.2022.54677

11. Yang H, Bhoo-Pathy N, Brand JS, Hedayati E, Grassmann F, Zeng E, Bergh J, Bian W, Ludvigsson JF, Hall P, et al. Risk of heart disease following treatment for breast cancer - results from a population-based cohort study. Elife. 2022;11. doi: 10.7554/eLife.71562

12. Paterson DI, Wiebe N, Cheung WY, Mackey JR, Pituskin E, Reiman A, Tonelli M. Incident Cardiovascular Disease Among Adults With Cancer: A Population-Based Cohort Study. JACC CardioOncol. 2022;4:85–94. doi: 10.1016/j.jaccao.2022.01.100

13. de Boer RA, Meijers WC, van der Meer P, van Veldhuisen DJ. Cancer and heart disease: associations and relations. Eur J Heart Fail. 2019;21:1515-1525. doi: 10.1002/ejhf.1539

14. Meijers WC, de Boer RA. Common risk factors for heart failure and cancer. Cardiovasc Res. 2019;115:844–853. doi: 10.1093/cvr/cvz035

15. de Boer RA, Hulot JS, Tocchetti CG, Aboumsallem JP, Ameri P, Anker SD, Bauersachs J, Bertero E, Coats AJS, Celutkiene J, et al. Common mechanistic pathways in cancer and heart failure. A scientific roadmap on behalf of the Translational Research Committee of the Heart Failure Association (HFA) of the European Society of Cardiology (ESC). Eur J Heart Fail. 2020;22:2272–2289. doi: 10.1002/ejhf.2029

16. Ahlers MJ, Lowery BD, Farber-Eger E, Wang TJ, Bradham W, Ormseth MJ, Chung CP, Stein CM, Gupta DK. Heart Failure Risk Associated With Rheumatoid Arthritis-Related Chronic Inflammation. J Am Heart Assoc. 2020;9:e014661. doi: 10.1161/JAHA.119.014661

17. Simon TA, Thompson A, Gandhi KK, Hochberg MC, Suissa S. Incidence of malignancy in adult patients with rheumatoid arthritis: a meta-analysis. Arthritis Res Ther. 2015;17:212. doi: 10.1186/s13075-015-0728-9

18. Codd V, Wang Q, Allara E, Musicha C, Kaptoge S, Stoma S, Jiang T, Hamby SE, Braund PS, Bountziouka V, et al. Polygenic basis and biomedical consequences of telomere length variation. Nat Genet. 2021;53:1425–1433. doi: 10.1038/s41588-021-00944-6

19. Codd V, Denniff M, Swinfield C, Warner SC, Papakonstantinou M, Sheth S, Nanus DE, Budgeon CA, Musicha C, Bountziouka V, et al. Measurement and initial characterization of leukocyte telomere length in 474,074 participants in UK Biobank. Nat Aging. 2022;2:170–179. doi: 10.1038/s43587-021-00166-9

20. Aviv A, Anderson JJ, Shay JW. Mutations, Cancer and the Telomere Length Paradox. Trends Cancer. 2017;3:253–258. doi: 10.1016/j.trecan.2017.02.005

21. Lin J, Epel E. Stress and telomere shortening: Insights from cellular mechanisms. Ageing Res Rev. 2022;73:101507. doi: 10.1016/j.arr.2021.101507

22. Epel ES, Blackburn EH, Lin J, Dhabhar FS, Adler NE, Morrow JD, Cawthon RM. Accelerated telomere shortening in response to life stress. Proc Natl Acad Sci U S A. 2004;101:17312–17315. doi: 10.1073/pnas.0407162101

23. Ahrens KA, Rossen LM, Simon AE. Relationship Between Mean Leucocyte Telomere Length and Measures of Allostatic Load in US Reproductive-Aged Women, NHANES 1999-2002. Paediatr Perinat Epidemiol. 2016;30:325–335. doi: 10.1111/ppe.12277

24. Zalli A, Carvalho LA, Lin J, Hamer M, Erusalimsky JD, Blackburn EH, Steptoe A. Shorter telomeres with high telomerase activity are associated with raised allostatic load and impoverished psychosocial resources. Proc Natl Acad Sci U S A. 2014;111:4519–4524. doi: 10.1073/pnas.1322145111

25. Bountziouka V, Hansell AL, Nelson CP, Codd V, Samani NJ. Large-Scale Analysis of the Association between Air Pollutants and Leucocyte Telomere Length in the UK Biobank. Environ Health Perspect. 2023;131:27701. doi: 10.1289/EHP11745

26. Zong ZQ, Chen SW, Wu Y, Gui SY, Zhang XJ, Hu CY. Ambient air pollution exposure and telomere length: a systematic review and meta-analysis. Public Health. 2023;215:42–55. doi: 10.1016/j.puhe.2022.11.022

27. Niehoff NM, Gammon MD, Keil AP, Nichols HB, Engel LS, Taylor JA, White AJ, Sandler DP. Hazardous air pollutants and telomere length in the Sister Study. Environ Epidemiol. 2019;3. doi: 10.1097/ee9.0000000000000053

28. Schmidt CW. Telomere Length and Air Pollution: Observations in Women Who Use Biomass Cookstoves. Environ Health Perspect. 2020;128:34005. doi: 10.1289/EHP6445

29. Factor-Litvak P, Susser E, Aviv A. Environmental Exposures, Telomere Length at Birth, and Disease Susceptibility in Later Life. JAMA Pediatr. 2017;171:1143–1144. doi: 10.1001/jamapediatrics.2017.3562

30. Wong JY, De Vivo I, Lin X, Christiani DC. Cumulative PM(2.5) exposure and telomere length in workers exposed to welding fumes. J Toxicol Environ Health A. 2014;77:441–455. doi: 10.1080/15287394.2013.875497

31. Aung N, Wang Q, van Duijvenboden S, Burns R, Stoma S, Raisi-Estabragh Z, Ahmet S, Allara E, Wood A, Di Angelantonio E, et al. Association of Longer Leukocyte Telomere Length With Cardiac Size, Function, and Heart Failure. JAMA Cardiology. 2023. doi: 10.1001/jamacardio.2023.2167

32. Xu C, Wang Z, Su X, Da M, Yang Z, Duan W, Mo X. Association between leucocyte telomere length and cardiovascular disease in a large general population in the United States. Sci Rep. 2020;10:80. doi: 10.1038/s41598-019-57050-1

33. Haycock PC, Heydon EE, Kaptoge S, Butterworth AS, Thompson A, Willeit P. Leucocyte telomere length and risk of cardiovascular disease: systematic review and meta-analysis. BMJ. 2014;349:g4227. doi: 10.1136/bmj.g4227

34. Telomeres Mendelian Randomization C, Haycock PC, Burgess S, Nounu A, Zheng J, Okoli GN, Bowden J, Wade KH, Timpson NJ, Evans DM, et al. Association Between Telomere Length and Risk of Cancer and Non-Neoplastic Diseases: A Mendelian Randomization Study. JAMA Oncol. 2017;3:636-651. doi: 10.1001/jamaoncol.2016.5945

35. Zhu X, Han W, Xue W, Zou Y, Xie C, Du J, Jin G. The association between telomere length and cancer risk in population studies. Sci Rep. 2016;6:22243. doi: 10.1038/srep22243

36. Gao Y, Wei Y, Zhou X, Huang S, Zhao H, Zeng P. Assessing the Relationship Between Leukocyte Telomere Length and Cancer Risk/Mortality in UK Biobank and TCGA Datasets With the Genetic Risk Score and Mendelian Randomization Approaches. Front Genet. 2020;11:583106. doi: 10.3389/fgene.2020.583106

37. Wentzensen IM, Mirabello L, Pfeiffer RM, Savage SA. The association of telomere length and cancer: a meta-analysis. Cancer Epidemiol Biomarkers Prev. 2011;20:1238–1250. doi: 10.1158/1055-9965.EPI-11-0005

38. Machiela MJ, Hsiung CA, Shu XO, Seow WJ, Wang Z, Matsuo K, Hong YC, Seow A, Wu C, Hosgood HD, 3rd, et al. Genetic variants associated with longer telomere length are associated with increased lung cancer risk among never-smoking women in Asia: a report from the female lung cancer consortium in Asia. Int J Cancer. 2015;137:311–319. doi: 10.1002/ijc.29393

39. Seow WJ, Cawthon RM, Purdue MP, Hu W, Gao YT, Huang WY, Weinstein SJ, Ji BT, Virtamo J, Hosgood HD, 3rd, et al. Telomere length in white blood cell DNA and lung cancer: a pooled analysis of three prospective cohorts. Cancer Res. 2014;74:4090–4098. doi: 10.1158/0008-5472.CAN-14-0459

40. Lan Q, Cawthon R, Gao Y, Hu W, Hosgood HD, 3rd, Barone-Adesi F, Ji BT, Bassig B, Chow WH, Shu X, et al. Longer telomere length in peripheral white blood cells is associated with risk of lung cancer and the rs2736100 (CLPTM1L-TERT) polymorphism in a prospective cohort study among women in China. PLoS One. 2013;8:e59230. doi: 10.1371/journal.pone.0059230

41. Liu M, Luo P, Liu L, Wei X, Bai X, Li J, Wu L, Luo M. Immune-mediated inflammatory diseases and leukocyte telomere length: A Mendelian randomization study. Front Genet. 2023;14:1129247. doi: 10.3389/fgene.2023.1129247

42. Thompson DJ, Wells D, Selzam S, Peneva I, Moore R, Sharp K, Tarran WA, Beard EJ, Riveros-Mckay F, Giner-Delgado C, et al. UK Biobank release and systematic evaluation of optimised polygenic risk scores for 53 diseases and quantitative traits. medRxiv. 2022:2022.2006.2016.22276246. doi: 10.1101/2022.06.16.22276246

43. Discacciati A, Bellavia A, Lee JJ, Mazumdar M, Valeri L. Med4way: a Stata command to investigate mediating and interactive mechanisms using the four-way effect decomposition. Int J Epidemiol. 2018. doi: 10.1093/ije/dyy236

44. Conrad N, Verbeke G, Molenberghs G, Goetschalckx L, Callender T, Cambridge G, Mason JC, Rahimi K, McMurray JJV, Verbakel JY. Autoimmune diseases and cardiovascular risk: a population-based study on 19 autoimmune diseases and 12 cardiovascular diseases in 22 million individuals in the UK. Lancet. 2022;400:733–743. doi: 10.1016/S0140-6736(22)01349-6

45. World Health Organization. https://www.whoint/data/gho/data/themes/mortality-and-global-health-estimates. 2023.

46. Ingebrigtsen TS, Marott JL, Vestbo J, Nordestgaard BG, Lange P. Coronary heart disease and heart failure in asthma, COPD and asthma-COPD overlap. BMJ Open Respir Res. 2020;7. doi: 10.1136/bmjresp-2019-000470

47. Sun D, Wang T, Heianza Y, Lv J, Han L, Rabito F, Kelly T, Li S, He J, Bazzano L, et al. A History of Asthma From Childhood and Left Ventricular Mass in Asymptomatic Young Adults: The Bogalusa Heart Study. JACC Heart Fail. 2017;5:497–504. doi: 10.1016/j.jchf.2017.03.009

48. Iribarren C, Tolstykh IV, Miller MK, Sobel E, Eisner MD. Adult asthma and risk of coronary heart disease, cerebrovascular disease, and heart failure: a prospective study of 2 matched cohorts. Am J Epidemiol. 2012;176:1014–1024. doi: 10.1093/aje/kws181

49. Chen H, Chen W, Zheng L. Genetic liability to asthma and risk of cardiovascular diseases: A Mendelian randomization study. Front Genet. 2022;13:879468. doi: 10.3389/fgene.2022.879468

50. Schneider CV, Schneider KM, Teumer A, Rudolph KL, Hartmann D, Rader DJ, Strnad P. Association of Telomere Length With Risk of Disease and Mortality. JAMA Intern Med. 2022;182:291–300. doi: 10.1001/jamainternmed.2021.7804

51. Deng Y, Li Q, Zhou F, Li G, Liu J, Lv J, Li L, Chang D. Telomere length and the risk of cardiovascular diseases: A Mendelian randomization study. Front Cardiovasc Med. 2022;9:1012615. doi: 10.3389/fcvm.2022.1012615

52. Zhan Y, Karlsson IK, Karlsson R, Tillander A, Reynolds CA, Pedersen NL, Hagg S. Exploring the Causal Pathway From Telomere Length to Coronary Heart Disease: A Network Mendelian Randomization Study. Circ Res. 2017;121:214–219. doi: 10.1161/CIRCRESAHA.116.310517

53. Prescott J, Wentzensen IM, Savage SA, De Vivo I. Epidemiologic evidence for a role of telomere dysfunction in cancer etiology. Mutat Res. 2012;730:75–84. doi: 10.1016/j.mrfmmm.2011.06.009

54. Ma H, Zhou Z, Wei S, Liu Z, Pooley KA, Dunning AM, Svenson U, Roos G, Hosgood HD, 3rd, Shen M, et al. Shortened telomere length is associated with increased risk of cancer: a meta-analysis. PLoS One. 2011;6:e20466. doi: 10.1371/journal.pone.0020466

55. McGrath M, Wong JY, Michaud D, Hunter DJ, De Vivo I. Telomere length, cigarette smoking, and bladder cancer risk in men and women. Cancer Epidemiol Biomarkers Prev. 2007;16:815–819. doi: 10.1158/1055-9965.EPI-06-0961

56. Risques RA, Vaughan TL, Li X, Odze RD, Blount PL, Ayub K, Gallaher JL, Reid BJ, Rabinovitch PS. Leukocyte telomere length predicts cancer risk in Barrett’s esophagus. Cancer Epidemiol Biomarkers Prev. 2007;16:2649–2655. doi: 10.1158/1055-9965.EPI-07-0624

57. Willeit P, Willeit J, Mayr A, Weger S, Oberhollenzer F, Brandstatter A, Kronenberg F, Kiechl S. Telomere length and risk of incident cancer and cancer mortality. JAMA. 2010;304:69–75. doi: 10.1001/jama.2010.897

58. Lan Q, Cawthon R, Shen M, Weinstein SJ, Virtamo J, Lim U, Hosgood HD, 3rd, Albanes D, Rothman N. A prospective study of telomere length measured by monochrome multiplex quantitative PCR and risk of non-Hodgkin lymphoma. Clin Cancer Res. 2009;15:7429–7433. doi: 10.1158/1078-0432.CCR-09-0845

59. Han J, Qureshi AA, Prescott J, Guo Q, Ye L, Hunter DJ, De Vivo I. A prospective study of telomere length and the risk of skin cancer. J Invest Dermatol. 2009;129:415–421. doi: 10.1038/jid.2008.238

60. Shen M, Cawthon R, Rothman N, Weinstein SJ, Virtamo J, Hosgood HD, 3rd, Hu W, Lim U, Albanes D, Lan Q. A prospective study of telomere length measured by monochrome multiplex quantitative PCR and risk of lung cancer. Lung Cancer. 2011;73:133–137. doi: 10.1016/j.lungcan.2010.11.009

61. Sanchez-Espiridion B, Chen M, Chang JY, Lu C, Chang DW, Roth JA, Wu X, Gu J. Telomere length in peripheral blood leukocytes and lung cancer risk: a large case-control study in Caucasians. Cancer Res. 2014;74:2476–2486. doi: 10.1158/0008-5472.CAN-13-2968

62. Machiela MJ, Lan Q, Slager SL, Vermeulen RC, Teras LR, Camp NJ, Cerhan JR, Spinelli JJ, Wang SS, Nieters A, et al. Genetically predicted longer telomere length is associated with increased risk of B-cell lymphoma subtypes. Hum Mol Genet. 2016;25:1663–1676. doi: 10.1093/hmg/ddw027

63. Wong JY-Y, Blechter B, Hubbard AK, Shi J, Hu W, Rahman M, Gadalla S, Machiela M, Rothman N, Lan Q. Abstract 3009: Measured and genetically-predicted leukocyte telomere length and lung cancer risk in the prospective UK Biobank. Cancer Research. 2023;83:3009–3009. doi: 10.1158/1538-7445.am2023-3009

64. Mamas MA, Sperrin M, Watson MC, Coutts A, Wilde K, Burton C, Kadam UT, Kwok CS, Clark AB, Murchie P, et al. Do patients have worse outcomes in heart failure than in cancer? A primary care-based cohort study with 10-year follow-up in Scotland. Eur J Heart Fail. 2017;19:1095–1104. doi: 10.1002/ejhf.822

65. Taylor CJ, Ordonez-Mena JM, Roalfe AK, Lay-Flurrie S, Jones NR, Marshall T, Hobbs FDR. Trends in survival after a diagnosis of heart failure in the United Kingdom 2000-2017: population based cohort study. BMJ. 2019;364:l223. doi: 10.1136/bmj.l223

66. Fry A, Littlejohns TJ, Sudlow C, Doherty N, Adamska L, Sprosen T, Collins R, Allen NE. Comparison of Sociodemographic and Health-Related Characteristics of UK Biobank Participants With Those of the General Population. Am J Epidemiol. 2017;186:1026–1034. doi: 10.1093/aje/kwx246

67. Sudlow C, Gallacher J, Allen N, Beral V, Burton P, Danesh J, Downey P, Elliott P, Green J, Landray M, et al. UK biobank: an open access resource for identifying the causes of a wide range of complex diseases of middle and old age. PLoS Med. 2015;12:e1001779. doi: 10.1371/journal.pmed.1001779

68. Codd V, Denniff M, Swinfield C, Warner SC, Papakonstantinou M, Sheth S, Nanus DE, Budgeon CA, Musicha C, Bountziouka V, et al. A major population resource of 474,074 participants in UK Biobank to investigate determinants and biomedical consequences of leukocyte telomere length. medRxiv. 2021:2021.2003.2018.21253457. doi: 10.1101/2021.03.18.21253457

69. Khera AV, Chaffin M, Zekavat SM, Collins RL, Roselli C, Natarajan P, Lichtman JH, D’Onofrio G, Mattera J, Dreyer R, et al. Whole-Genome Sequencing to Characterize Monogenic and Polygenic Contributions in Patients Hospitalized With Early-Onset Myocardial Infarction. Circulation. 2019;139:1593–1602. doi: 10.1161/CIRCULATIONAHA.118.035658

70. Wong JYY, Bassig BA, Loftfield E, Hu W, Freedman ND, Ji BT, Elliott P, Silverman DT, Chanock SJ, Rothman N, et al. White Blood Cell Count and Risk of Incident Lung Cancer in the UK Biobank. JNCI Cancer Spectr. 2020;4:pkz102. doi: 10.1093/jncics/pkz102

71. Wong JYY, Jones RR, Breeze C, Blechter B, Rothman N, Hu W, Ji BT, Bassig BA, Silverman DT, Lan Q. Commute patterns, residential traffic-related air pollution, and lung cancer risk in the prospective UK Biobank cohort study. Environ Int. 2021;155:106698. doi: 10.1016/j.envint.2021.106698

